# Lived experience of people living with Podoconiosis in Southern Ethiopia: a phenomenological study

**DOI:** 10.1101/2025.10.17.25338075

**Authors:** Muluken Mussie Bibbiso, Yohannes Kebede, Fitsum Nigatu, Esayas Tadele

**Affiliations:** School of Public health, College of Health Science and Medicine, Wolaita Sodo University, Wolaita Sodo, Ethiopia; Department of Health, Behavior and Society, Faculty of Public Health, Jimma University, Jimma, Ethiopia; Department of Public health, College of Health Science and Medicine, Mettu University, Mettu, Ethiopia

**Keywords:** Key terms: Podoconiosis, lived experience, Southern Ethiopia

## Abstract

**Background:** Podoconiosis is a neglected tropical disease (NTD) that causes lower-leg swelling (lymphedema) and is caused by prolonged contact with irritating red-clay soil. Podoconiosis affects 4 million people worldwide, with Ethiopia having the highest prevalence. The disease has significantly increased the social, psychological, and financial strain on those who are affected. However, few studies have explored the real-world experiences of people living with Podoconiosis in Ethiopia. Therefore, this study aimed to explore the lived experiences of people living with Podoconiosis in Southern Ethiopia.

**Methodology:** A descriptive phenomenological study design was carried out to explore the lived experiences of 14 purposively selected people living with Podoconiosis in Sodo Zuriya district, Wolaita zone, Southern Ethiopia. In-depth interviews were conducted in the local language using an interview guide. All interviews were recorded using a digital audio recorder. The collected data were transcribed and translated to English and the data were analyzed using the qualitative data management software Atlas-ti version 7.5.16. A thematic analysis approach was used for analyzing the data and reporting the results.

**Results:** The thematic analysis of data revealed six prominent themes. The perceived cause of Podoconiosis, socio-economic impact, physical health experiences, psychological health problems, health care experiences, and coping mechanisms. The participants perceived that the causes of Podoconiosis were action of witchcraft, heredity, walking barefoot, and a curse from God or an evil spirit. As a result of Podoconiosis, patients suffered from difficulty walking, pain, wound development, and an associated bad odor, were banned by their family and community members, and felt hopeless. Patients used different coping mechanisms, including social support, traditional medicines and plants, they also expressed frequent washing of feet.

**Conclusion:** People living with Podoconiosis experienced social, physical, and psychological health problems such as stigma, isolation, fear, shame, and activity limitations. Healthcare organizations should provide integrated services for people living with Podoconiosis, including physical therapy, psychological support, social reintegration, and available services for Podoconiosis patients in health centers.

## Background

Podoconiosis (endemic non-filarial elephantiasis) is a chronic non-infectious neglected tropical disease (NTD) which affects the lower limb (1). Although the causes of Podoconiosis is not fully understood, current evidence suggest that it is caused by longstanding exposure to red clay soil of volcanic origin (2). It is believed that mineral particles that are absorbed via the skin of the foot are taken up by macrophages in the lower limb lymphatics and cause an inflammatory reaction that results in fibrosis and obstruction of the vessel lumen (3).

There are 4 million people worldwide who have Podoconiosis, mostly in tropical Africa, Central and South America, and Southeast Asia (4). Podoconiosis has been found to exist in 32 different countries. The global prevalence of Podoconiosis varies from 8.08% in Cameroon to 2.51% in Tanzania (1).The greatest burden of Podoconiosis globally is assumed to occur in Ethiopia (5). The disease is common in Ethiopia with more than 345 endemic disease-related woreda (6). In areas with irritant soil, the prevalence of Podoconiosis is approximately 5%-10% (7). Other studies have estimated that as many as 1.5 million people living with Podoconiosis in Ethiopia with 35 million people at risk of contracting a disease in the state (8).

The national average prevalence was 4% with the highest prevalence in the Southern Nations Nationality and People Regional (SNNPR) (8.3%) followed by Oromia (4%) and Amhara (3.9%) regional states (9). Most studies revealed the prevalence was even higher among those aged 15– 64 years who were economically active even one study confirmed that 64% of Podoconiosis cases are in the economically productive age group (7,8). Podoconiosis’ patients lose approximately half of their total number of productive workdays. The direct and productivity costs of Podoconiosis have been estimated at US$16 million per year in a population of 1.5 million people, imposing an economic burden of $208 million per year in Ethiopia (10).

The disease has significantly increased the social, psychological, and financial strain on those who are affected (11). Podoconiosis hence poses a serious danger to educational and employment opportunities (12). These patients also suffer from severe physical impairment, psychological comorbidity, and recurrent episodes of systemic sickness as a result of acute Adenolymphangits attacks (ALAs) (13). Several studies revealed that patients with Podoconiosis disease had a lower quality of life (14–16).

As documented in previous studies, people with lower limb lymphedema, such as Podoconiosis, were more likely to develop mental disorders than healthy people, suggesting that the psychological and social aspects of their quality of life were worsened (16). The presence of this disease has a detrimental effect on people’s quality of life. Patients with Podoconiosis experience a decrease in their physical, psychological, environmental, and social quality of life (14,16).

Evidence from Ethiopia demonstrated that patients’ psychological and social health were significantly impacted by social stigma related to Podoconiosis (14). The condition makes people and their families more likely to be stigmatized by their neighbors, making it one of the most common causes of social exclusion. Patients are banned from social events like weddings, funerals, schools, local meetings, and churches (11,17). A study conducted in Southern Ethiopia revealed that Podoconiosis patients faced stigma in their daily interactions. Unwillingness to marry with Podoconiosis patients, avoiding physical contact, spitting on patients, pinching nose when walking past patients at a distance were mentioned by patients as a manifestations of stigma (18). Most patients described it as ‘the worst disease’ mainly due to its negative social and psychological consequences (19).

Evidence has shown that one–sixth of the world’s population, mostly in developing countries, is infected with one or more NTDs (20). The WHO identified twenty NTDs for control and elimination at the global level by 2030, and among these diseases, nine were identified, including Podoconiosis, as a priority in Ethiopia with a range of endemicity across the regions. The third national NTD strategic plan 2021-2025 targets to eliminating and controlling Podoconiosis in all endemic woreda through lymphedema management, psychosocial support, social behavioral change communication and integration with Water, sanitation and hygiene(WASH) advocacy. The Strategic Plan is also aligned with the overall Health Sector Transformation Plan framework, HSTP II and the new WHO NTD Road map (2021–2030). Above all, exploring the lived experiences of people living with Podoconiosis has great value for resolving the global and national burdens of Podoconiosis. People living with Podoconiosis need the promotion of public and professional education about their condition to improve their quality of life (21–24).

Despite the significant health burden of Podoconiosis in Ethiopia, there is a critical evidence gap regarding the lived experiences of affected individuals. Most existing studies focus on prevalence, risk factors, geographical distribution and clinical aspects, neglecting qualitative insights into personal narratives and emotional struggles. Cultural perceptions and stigma surrounding Podoconiosis remain underexplored, limiting understanding of community perception towards the disease. Additionally, the economic impact on livelihoods and barriers to accessing healthcare services require further investigation. The psychological effects, including anxiety and social isolation, have not been adequately addressed. Furthermore, existing literature often overlooks the coping mechanisms and resilience strategies employed by individuals. Linking personal experiences to policy implications can enhance public health responses, yet this connection is largely absent in existing literatures. This shortage of information regarding the issue of Podoconiosis in Ethiopia particularly in the Southern Ehiopia, may hinder efforts to address the psychosocial, physical, economic and health care related consequences of Podoconiosis. There is a critical need for qualitative research that captures the voices of affected individuals, exploring their daily struggles, perceptions, and the impact on their livelihoods. Understanding these experiences can inform more effective interventions and policies tailored to support this marginalized population. Addressing this gap will contribute to a holistic understanding of Podoconiosis and enhance advocacy efforts for better healthcare access and social support systems.

## Methods and materials

### Study design and Setting

A descriptive phenomenological study design was conducted in Sodo Zuriya district Wolaita Zone, Southern Ethiopia. Sodo Zuriya Woreda is one of 16 rural Woreda and 6 town administrations in Wolaita zone. The administrative seat of Sodo Zuriya Woreda is Sodo town, which is located 383 km far from Addis Ababa (the capital city of Ethiopia) (25). Based on the 2019 population projection conducted by the Central statistical agency (CSA), this woreda has a total population of 200,911, of whom 98,824 are men and 102,087 women (26). The majority of people in the region earn their livelihood from subsistence farming. Farmers in the study area rarely wear shoes while working in their fields and hence are in direct contact with red clay soil. Direct and indirect contact of subsistence farmers with the red clay soil leads to a high prevalence of non-filarial elephantiasis (Podoconiosis) in the area. According to Federal Ministry of Health report (FMOH) Sodo Zuriya district is one of 15 districts in Wolaita Zone with high prevalence of Podoconiosis (18,27). In the district, there are thirty-seven health posts and six health centers. Additionally, there is Mossy Foot International, an organization which offers prevention and treatment of Podoconiosis. The study was conducted from June 1 to June 30, 2023.

### Study Participants

The study participants were purposively selected Podoconiosis patient who were clinically diagnosed and registered in Sodo Zuriya district. Fourteen Podoconiosis patients in Sodo Zuriya district participated in this study. New participants were approached until data saturation had been reached (until no new ideas were emerging).

### Sampling Technique

The participants were selected using the purposive sampling technique. Purposive sampling enables researchers to select participants with in-depth knowledge of the topic, facilitating the collection of rich, and detailed information. This approach ensures that the sample size achieves conceptual saturation, enhancing the researchers’ understanding of the issue (28). The criteria for selecting participants includes: clinically diagnosed and registered in Sodo Zuriya district, age >18 years. Participants were recruited with the help of health extension workers and a neglected tropical disease focal person in the woreda health office.

### Data Collection Method and Procedure

Data collection was carried out using in-depth interviews. An interview guide was developed based on the literature review and the investigator reflection on Podoconiosis patient lived experiences **(Additional file 1)**. The same topic guide was used for all participants to allow flexibility for any additional issues to be discussed. After establishing contact, the scope and objectives of the study were explained to the potential participants. After obtaining the necessary information from the registration books from the woreda health office, health extension workers invited the study participants who fulfilled criteria for interviews, and the interviews were conducted home to home visit. In depth interviews were conducted one-to-one in a quiet and private place to respect patient confidentiality. The time of interview was scheduled based on mutual agreement with the participants. In-depth interviews were carried out with Podoconiosis patient, utilizing a team of two experienced researchers who possess a strong background in qualitative data collection. The principal investigator (PI) played a crucial role as both a data collector and facilitator, ensuring the smooth progress of the interviews. To maintain objectivity and minimize potential biases, the PI had no prior connections or relationships with the research participants. The in-depth interview guide was prepared first in English and then translated into Wolaita (local language in the community) and retranslated back to English to check for consistency by professional and experienced translators, while upholding the integrity and fidelity of the data. All participants agreed to be audio-taped, and interviews lasted between 45 and 92 min. Data were obtained in the form of digital audio records, and the participants’ emotions and other nonverbal communication were recorded as field notes. During the interview, the researcher asked questions, took notes, and recorded participants’ non-verbal expressions. Data collection and analysis occurred simultaneously. Audio recordings of the interview and the field notes were transcribed.

### Data processing and Analysis

Thematic analysis approach was used for data analysis. The recorded material was transcribed verbatim, including emotional expressions and translated into English by professional and experienced translators. The transcribed data were exported to Atlas ti version 7.5.16 software for analysis. Transcripts were read several times to acquire a better grasp of the data and meaning units. Next up, Research team coded (highlighted sections of text – usually phrases or sentences – and come up with shorthand labels or “codes” to describe their content using the software). The coding started from the richer document and Research team went through the transcript of each interview. Then, highlighted all the phrases and sentences that match the previous codes and kept on adding new codes as go through the transcripts. After going through the codes, Research team organized the codes into categories. Then, looked over the codes and categories which had created, identified patterns among them, and started coming up with themes. To make sure that themes are useful and accurate representations of the data, returned to the data set and compared themes against it. Finally, the codes were sorted into relevant categories, and the main themes and categories were identified **(Additional file 2)**. Significant quotations were clustered into themes, and then used to explain participant experiences and the context that influenced how the participants experienced the phenomenon.

### Trustworthiness

To ensure the rigor and trustworthiness of the study, the principles of credibility, transferability, dependability, and confirmability were maintained. Credibility was enhanced through prolonged engagement, multiple interviews, triangulation of audio recordings and field notes, participant validation, and supervision by qualitative research experts. Transferability was supported by purposive sampling, detailed descriptions of the study context and recruitment methods, and the use of direct quotations to illustrate themes. Dependability was ensured through transparent documentation of data collection and analysis procedures, with audits by advisors and experts to verify consistency. Confirmability was achieved through peer review, expert verification, and maintaining an explicit audit trail to demonstrate that findings were grounded in the data.

## Results

### Socio-demographic characteristics of the participants

In this study, 14 in-depth interviews were conducted with Podoconiosis patients who live in Sodo Zuriya district, Wolaita Zone (Table 1

**Table 1:** Socio-demographic characteristics of participants in Sodo Zuriya district, Wolaita Zone Southern Ethiopia, 2023.

| Variables | Classifications | Frequency |
| --- | --- | --- |
| Age | 15-24 | 1 |
|  | 25-34 | 3 |
|  | 35-44 | 6 |
|  | 45+ | 4 |
| Sex | Male | 9 |
|  | Female | 5 |
| Educational status | Illiterate | 6 |
|  | Elementary and secondary | 8 |
| Current marital status | Single | 1 |
|  | Married | 9 |
|  | Widowed | 3 |
|  | Divorced | 1 |
| Religion | Orthodox | 6 |
|  | Protestant | 8 |
| Occupation | Farmer | 10 |
|  | Merchant | 2 |
|  | Housewife | 2 |
| How long have you been living with Podoconiosis? | 1-10 | 2 |
|  | 11-20 | 5 |
|  | 21-30 | 4 |
|  | 30+ | 3 |
| Residence | Rural | 14 |

### Theme 1: Perceived cause of Podoconiosis

#### Action of witchcraft

The action of witchcraft in the context of this study refers to the beliefs or perceptions held by participants that this Podoconiosis is caused by witchcraft practices. Participants expressed that contact with a witchcraft or sorcery was a contributing factor to Podoconiosis. People in the community may not have access to accurate medical information about the causes of Podoconiosis, leading them to attribute this to the condition of the witchcraft. In areas where belief in witchcraft is prevalent, traditional healers may be consulted to treat Podoconiosis.

One of the respondents stated,

> *“I got this disease because of witchcraft. Because one day I was exposed to dead sheep’s blood on the road, I touched the blood in my right leg. This is witchcraft, because my leg swelled. Therefore, in my view, Podoconiosis is a disease that affects the lower leg and is caused by witchcraft.”* (IDI, 10)

> Strengthen by saying, *“I don’t exactly know this disease, but in my view, Podoconiosis is a disease caused by witchcraft. Three of my children passed away due to witchcraft. After my children’s deaths, I was exposed to witchcraft because I walked barefoot daily and was exposed to cold weather conditions.”* (IDI, 8)

#### Heredity

Heredity in the context of this study refers to the beliefs or perceptions held by participants that this Podoconiosis cause is related to hereditary or generational line. Some of the participants verbalized that heredity was the cause of Podoconiosis. The quotes below highlights further

> *“I’ve seen my father and grandfather suffer from swollen legs just like mine. It feels like this condition runs in our family, like it’s something we inherit.“* (IDI, 9)

> *“My mother always told me that our family has a history of swollen legs. It seems like every generation has faced this struggle. I can’t help but think that it’s something passed down through our blood. When I look at my own children, I worry they might inherit this burden too.”* (IDI, 2)

#### Walking barefoot

It became evident in the data that other participants perceived walking barefoot as the cause of Podoconiosis. As per the idea obtained from participants, they believed that the cause of Podoconiosis was walking barefoot since this patient’s source of information was health professionals. They attended health facilities which offer services for Podoconiosis. One of the study participant well mentioned as follows:

> *“In my view, the cause of this swelling might be not wearing shoes and not keeping the feet clean. I wore no shoes. Not just me, but the rest of my family didn’t wear shoes, which may have contributed to the illness. When we go barefoot, we could come into contact with various worms and other chemicals on the ground that could result in swelling. I still don’t wear shoes when I harvest my land since it doesn’t make me feel comfortable.”* (IDI, 11)

Also, participants reported that barefoot exposure to poison and worms was perceived as a cause of Podoconiosis. Although the term soil or dust was rarely mentioned, participants referred to poison and other harmful things in the soil. Because of walking barefoot, the feet trap dust through sweating, and foot swelling develops consequently. Because of walking barefoot, worms in the soil enter the feet and cause swelling. A verbatim quotation obtained from a study participant well describes this result:

> *“In my view, the cause of this condition is exposure to poison. Because of walking barefoot, the feet trap dust through sweating, and foot swelling consequently develops. Because of walking barefoot, worms in the soil enter the feet and cause swelling.”* (IDI, 3)

According to this study, a curse from God or evil spirt implies a supernatural punishment or affliction imposed by a deity, specifically God, on an individual or a group of people for their sins or family’s sin or disobedience. Participants noted that Podoconiosis is caused by a curse from God or an evil spirt. This notion is well explained as follows;

> *“Podoconiosis is a frustrating and boring disease that I got due to being shunned from the community or a curse from God or an evil spirit.”* (IDI, 4)

### Theme 2: Socio-economic impact

Socio-economic impact is the second major theme inductively developed from the participant responses. This theme developed from participant responses that explain experiences that can have a significant impact on the daily lives of individuals with Podoconiosis, making it difficult for them to work, engage in relevant activities of daily living, and participate in social activities. The theme is comprised of two basic sub-themes, which are social impact, and economic impact.

#### Social impact

##### Difficulty finding a marriage partner

This study suggested that one of the difficulties Podoconiosis patients face is finding a partner for marriage for those who are in need of marriage and for their children to marry to non-patient families. Many people believe that people affected by Podoconiosis, including their family members, should not have a loving relationship, get married or have a child, and some think that the question of marriage relationships is disgusting. In this study, participants experienced difficulty finding a marriage partner because healthy individuals do not allow their children to marry individuals suffering from Podoconiosis. A female participant explained this result in the following manner:

> *“Since no one prepares patients with Podoconiosis for marriage, the disease has a bad reputation in our community. The families of healthy people forbade their children from getting married to people who had Podoconiosis. Because people in our society think that this disease is genetic and contagious, I had trouble finding a spouse. Due to their good health, all of my friends were married between the ages of 16 and 18. When I was at age of 20’s, a man outside of our Kebele prepared me for marriage, and we intended to live together. However, later that year, someone in my neighborhood told him I suffered from leg swelling, and he refused to marry me.”* (IDI, 11)

One male Podoconiosis patient added by saying like this,

> *“Most often, I stayed idle and was disregarded. I have faced difficulty finding a marriage partner due to this condition. In our community, people believe that this disease is a curse from God and an evil spirit. I tried more than three times to find a marriage partner, all of which failed due to this boring disease.”* (IDI, 4)

##### Divorce

According to this study, the patients also have a high probability of divorcing their spouse after developing the disease. The unaffected partner does not want to continue living with the affected individual under the same roof. The experience of one woman explains this phenomenon best:

> *” …… It [Podoconiosis] has negatively affected my social life. Podoconiosis is aggravated by long walks and hard work. Due to this disease, my husband threw me out (divorced), and I am living alone.”* (Feeling sad). (IDI, 2)

> *“A year goes by during which my husband sleeps in a different bed at the same house; especially during acute attacks, he doesn’t even want to sit with me. After some time, he believed I would get better. He had given up on me, though, and had repeatedly considered divorcing me, but his father had opposed him. If you become as useless as I am, nobody will want you.”* (IDI, 1)

##### Difficulty participating in social activities

The study participants revealed that one of the social problems they faced was their inability to participate in various social activities in the community. This resulted from either the belief from the patients’ side that the unaffected members of the community stigmatize or discriminate them because of their disease or from the negative attitude of the unaffected community members towards Podoconiosis patients. A participant who had lived with Podoconiosis for the last 23 years explained as follows:

> *“……I asked my God what had been given to me and how I could be healed of this illness. I can’t hang around with my friends for very long. I am unable to freely engage in social activities. I am unable to sit with others during the coffee ceremony because I am afraid and ashamed. So why do I live? People cover their faces and mouths with towels and their hands when I approach them.”* (IDI, 12)

Some participants prefer to stay at home because people in their community believe that Podoconiosis is contagious and a curse from God. A participant responded:

> *“If there are no urgent matters, I prefer staying at home. From my experience, it is better to not make contact with people. People surrounding us expressly avoid us, and they continually label us patients with a contagious and boring disease.”* (IDI, 3)

##### Discrimination and stigma

The majority of participants reported that they were the target of discrimination by families, neighbors, community members, and other social groups due to Podoconiosis and the bad odor which was more aggravated during acute attacks, and they also reported insults and isolation due to Podoconiosis. This notion is well explained by participants as follows:

> *“My mother passed away a year after I developed feet swelling, and my father then married another woman. As you know, stepmothers can be cruel to their husband’s children, even if they are healthy. My stepmother often called me “Geddiya Kixxe.“(Means mossy foot in English) She didn’t treat me well. She didn’t even want to mention me to the family or have a meal with me.”* (Feeling sad). (IDI, 12)

> *“While I was a student, most of my classmates would not accept me sitting near them because of my condition. Actually, not only at school. Generally, people would stand up and leave me isolated whenever I came to sit near them.”* (IDI, 6)

##### Insult

Some participants experienced insults due to the illness; community members and family members teased and insulted Podoconiosis patients; they considered Podoconiosis patients different and feared the condition was transmitted and contagious. Therefore, they don’t want to sit with them for a long period of time. This notion is well explained as follows;

> *“I have been teased and insulted by most of my friends, including my brother. They consider me different and they fear that my condition [Podoconiosis] will be transferred to them if they sit with me and stay with me for a long time. Sometimes community members, especially neighbors, didn’t invite me to any ceremonies after I was ill because they [my family and neighbors] didn’t remember me; they thought I might have the illness due to the severe pain and bad odor of my feet.”* (IDI, 10)

> *“The people belittle me and call me the woman with mossy feet “Geddiya Kixxa”. I am bullied by my neighbors because of my legs. When I go to weddings and funerals, people spit on me and insult me. I feel very sad.”* (Crying hanging down to ground) (IDI, 2)

#### Economic impact

##### Difficulty of work engagement

According to the participant’s responses, the Podoconiosis patient’s lower limbs were impaired, and walking and standing for extended periods of time was associated with difficulty; this could make the patient dependent on others and unable to work. This difficulty was also a barrier to them engaging in relevant activities to the best of their abilities, and preventing them from integrating economically into the community. In other words, they cannot perform what they are supposed to perform at an appropriate time and place compared to unaffected community members. This result is well captured by participant in the following ways:

> *“This illness made me less productive. I can’t farm because of the physical challenges brought on by the sickness; therefore, I’m doomed to be poor. If you are unable to properly care for your land and engage in farming, your only option is to beg for food. I believe I am destined to be impoverished because of this illness.”* (IDI, 13)

Another women added;

> *“I wish I could work, but because of this illness, I am unable to do so. My social life has been wrecked by it. Podoconiosis is made worse by strenuous activity. It is a dull illness; consider not going to work at your office regularly and not getting paid; my situation is similar to this. Due to this illness, I am living the worst life among the residents of our community.”* (IDI, 12)

##### Being dependent on others

In the context of this study, being dependent on others refers to relying on other individuals (families) or entities for various aspects of one’s life, such as financial assistance and physical support. This dependency can arise due to a variety of factors, including activity limitations, physical disability and pain. It involves an individual’s inability to fully meet their own needs without the assistance or involvement of others. Participants described that they were dependent on others due to inability to perform daily activities. They beg their families for food. One of the study participant said;

> *“Economically, I am dependent on my husband. I am not capable of doing work, and as a result, I am leading a lower lifestyle. Even though I was making an effort to tolerate working, the pain in my lower leg hindered me from undertaking my routine activities at home and outside. I have also stopped making pottery to sell, which lowered my income. I have nothing and lead a miserable life”* (IDI, 1)

##### Losing productivity

According to participants’ responses, the illness reduced the energy and ability to work which reduced work hours and increased work absents and led to loss of productivity. Additionally, participants reported that they lead miserable life due to the illness. The illness is well aggravated during acute attack. During acute attacks, participants reported that they were bedridden for a long period of time and were not able to perform their daily activities, which led to loss of productivity. This result is well explained as follows;

> *“This disease reduced the energy with which I could work, reduced my working hours, increased work absenteeism, and decreased my productivity. I could not earn as much income as Podoconiosis free individuals in our community.”* (IDI, 7)

Another participant also added;

> *“During the acute attack, I couldn’t work as a result of the disease. I have nothing and lead a miserable life because I lost my productivity. When I started to work on my every day activities, like farming, the condition aggravated, and I felt sick. The last week with my elder child, we started to harvest maize and teff but I faced difficulty working due to severe pain in my feet.”* (IDI, 6)

### Theme 3: Physical health experiences

The physical health experience is very evident to victims because of the manifestations of swelling, especially in the lower extremities, making it more difficult for them to move around and perform their activities of daily living. Most of the participants experienced difficulty walking/activity limitations, and an undesirable odor. Because of these manifestations, they are often not able to walk long distances or cannot walk on the farm, thus making life a day-to-day struggle.

#### Activity limitation/Inability to walk

People living with Podoconiosis often experience physical activity limitations due to swelling and discomfort in their lower extremities. This condition can lead to pain, stiffness, and reduced mobility, making it challenging for affected individuals to engage in activities that require standing or walking for extended periods. As a result, they may have difficulty performing daily tasks such as walking long distances, standing for prolonged periods, or engaging in agricultural work, which can significantly impact their quality of life. Most of the study participants described their inability to walk and their activity limitations. This notion is well explained as follows:

> *“When I was healthy, I could do everything I wanted to, but ever since this condition entered my life, I have had trouble leading a joyful existence. For instance, this condition affects my physical health and makes it difficult for me to walk long distances.”* (IDI, 13)

Another respondent also supports, “*I can’t stretch my leg; it is difficult for me to walk. I am bedridden because the disease is so painful. No one understood what was happening with my feet.*” (IDI, 2)

#### Bad odor

Participants reported that they experienced a bad odor in their feet, which was more aggravated during the acute attack. One of the physical health problems (challenges) that Podoconiosis patients’ face in integrating with the general community is the bad smell of their feet. According to this study, the stinky smell of their feet forced them to isolate themselves from social gatherings and various ceremonies. A Podoconiosis patient described the following:

> *“As you can see now, it is very damaged and disfiguring. I do not even sit for long periods of time with people because of the smell; it has a bad odor or foul smelling and because of the wound, flies bother me.”* (IDI, 1)

Additionally, a woman with Podoconiosis added:

> *“Since my disease has now progressed and my feet smell horrible, I might avoid going to various social events like weddings and funerals.”* (IDI, 14)

#### Burning pain

The burning pain experienced by individuals with Podoconiosis can significantly impact their quality of life. It can hinder their ability to perform daily activities, leading to further complications and social isolation. Participants experienced severe pain, which was more aggravated during acute attacks and aggravated by cold weather conditions and the rainy season. This is well explained as follows;

> *“I had one experience where I was at a funeral ceremony that was far away from my home. When the programme was over, everyone got up to go to their houses, but I couldn’t because the pain in my leg prevented me from walking with them. I was starting to leave slowly when they left.”* (IDI, 9)

### Theme 4: Psychological health problems

#### Depression

The study participants reported feeling depressed and sad that they were not involved in socio-cultural activities and preferred to be alone sometimes. The majority of participants described being depressed at some point. Participants expressed a preference for being alone instead of being with their community members and families, fearing that they might disturb people. They reported that they became sad when they received negative responses from others. A participant who had been living with Podoconiosis for the last 31 years shared his story as follows;

> *“……. As I told you earlier, I am in a painful condition since I lost my vision and am leading a boring life. My relatives and neighbors treat me badly. They don’t look at me because of this disease. I feel bad because they don’t treat me the way they did before. They don’t help me. I think they are afraid or repulsed. Because of this disease, they don’t love me. I feel sad. I don’t go out unless it’s really necessary. I am always locked up. I don’t like people looking at my legs. I feel very depressed and sad. I am alone, and I don’t spend time with anyone anymore. I couldn’t see anything. I lost my vision, not only my leg. I feel guilty and depressed. I blame God, why did he give me this? During an acute attack, everything changes to darkness, and no one helps me since I am blind.”* (Crying) (IDI, 9)

> *“On Sundays, I would lie in bed crying as I listened to the sound of people singing at church services I could not attend, because of the disfiguring appearance and bad odor of this disease. I feel depressed and sad.”* (IDI, 1)

#### Feeling hopeless

Participants discussed that their situation was so psychologically distressing that they felt hopeless and described great despair. Participants think about their future and feel hopeless since the disease has made them bedridden. Participant said;

> *“However, when the condition develops, I am confined to bed. I usually worry about my feet when I’m sick (have an acute illness), as I can’t keep up with my other healthy buddies or friends. Therefore, it has a major influence on me when I am sick. I become frustrated that I cannot be as healthy as my friends.. I don’t have hope that I will be better when the attack happens to me.”* (IDI, 12)

#### Suicidal thoughts

In the context of this study, suicidal thoughts refer to the presence of persistent thoughts about ending one’s own life due to Podoconiosis. These thoughts can range from fleeting considerations of suicide to more detailed and specific plans for self-harm or suicide. Participants had a history of suicide attempts due to Podoconiosis negative social and psychological consequences. The participants described their thoughts of wishing they were dead. A participant said;

> *“You are aware of how awful it is to have this offensive scent and disfiguring appearance around your family and neighbors. It would be better if I passed away. I frequently prayed to God and said that I preferred death. I am the unluckiest person in this world.”* (IDI, 9)

Another Podoconiosis patient also mentioned;

> *“I feel worried because it [Podoconiosis] prevents me from working. I always wonder why this happened to me. Why I am usually unable to work hard and why I am inferior to my relatives and friends? Why did the Almighty God only make me ill? I always think badly about the future, and sometimes I feel like killing myself [suicidal ideation]. I was thinking that I shouldn’t be alive as an inferior human being. I had to take some toxins and die. (Her eyes were filled with tears.) After a while, I improved to some extent; at least I could walk, eat, and drink following the treatment. Even I cannot sit close to my mother and father. I tried to kill myself several times.”* (Crying…) (IDI, 1)

### Theme 5: Healthcare-related experiences

Healthcare related experience is the fifth major theme inductively developed from participant responses. This theme developed from participant responses that explain the interactions and encounters people living with Podoconiosis have within the healthcare system, including their interactions with healthcare providers, the quality of care received, and the overall impact on their well-being. These experiences encompass a wide range of factors, such as access to care, communication with healthcare professionals, coordination of services, and the physical environment of healthcare facilities.

#### Access to health care related experiences

In the context of this study, access to health care related experiences refers to the experiences of people living with Podoconiosis to obtain, and use needed health services. It encompasses a wide range of factors, including physical accessibility (distance), affordability, and availability of services. Participants noted that they travelled long distance to obtain services, which can be a significant barrier to obtaining timely and appropriate health care.

> *“I went to Wolaita Sodo Otona and Gununo (nearby towns and capital cities of Boloso Bombe Woreda) to seek care. I have been taking treatment from the Otona Podoconiosis treatment and prevention center, because there are no services provided for Podoconiosis patients in our health center.”* (IDI, 2)

The unavailability of services as experienced by people living with Podoconiosis refers to the situation in which patients are unable to access the healthcare services they need. The experience of unavailability of services can have significant implications for patients, leading to delays in treatment, exacerbation of health conditions, increased stress and anxiety, and overall dissatisfaction with the healthcare system. Participants discussed that they went to their nearby health center to seek care, but the unavailability of services in the center prevented them from receiving services. In the majority of areas where Podoconiosis is prevalent, access to healthcare, including physical health services, mental health services, and counselling is limited. Patients may not receive the necessary support and treatment for their physical and mental health needs. For instances, a woman stated this result as follows;

> *“……. However, there is no service provided for Podoconiosis patients in our health center. Sometimes I wondered why the government paid little attention to Podoconiosis patients. I went to our nearby health center to seek care, but they only offered some painkillers. There is no service provided for Podoconiosis patients in our health center.”* (IDI, 12)

#### Patient-provider relationship

In the context of this study, the patient-provider relationship refers to the interaction and connection between a patient and their healthcare provider. This relationship is fundamental to the delivery of healthcare services and encompasses various aspects such as communication, trust, empathy, shared decision-making, and mutual respect. According to respondents’ responses, poor patient-professional relationships and negative perceptions of health professionals, for instance, treating Podoconiosis patients badly was reported. For instance, a man stated this result as follows;

> *” However, health professionals in our health center don’t treat us politely. I recall it being a dreadful experience being insulted by a health professional. When I talked about my illness to the health professionals, they said that there is no treatment or service for this disease in our health center.”* (IDI, 8)

On the other hand, some of the participants reported that they received good services and that the health professionals’ approach was good for them. On the positive side of the health professional reaction, the patients were satisfied with health professional as they improved.

One respondent said;

> *“My neighbor affected by Podoconiosis told me about Mossy Foot International (Non-Governmental organization which offers services for Podoconiosis patients in the Wolaita Zone), which is found in Sodo and offers treatment and advice related to the disease. Some kind of hope started to shine inside me. The next morning, I went there to get treatment. When I reached there, all the staff cooperated and gave me bandages, bleach, ointment, and Whitefield. Health professionals in the center treat me well.”* (IDI, 7)

### Theme 6: Coping mechanisms for Podoconiosis

Coping mechanisms for Podoconiosis is the sixth major theme inductively developed from participant responses. This theme developed from participant responses that explains a variety of coping mechanisms to deal with physical health problems, social health problems, and psychological health problems. The theme is comprises of five basic sub-themes:

#### Religious institutions and praying to God as a coping strategy

According to this study, participants mentioned religious institutions (holy water) and praying to God as coping mechanisms. Praying and performing religious rituals help the patients manage their stress. Some Podoconiosis patients are devoted to their religion and have a strong belief in God. They went to religious institutions and used holy water as a treatment option. In addition, they also got psychological as well as spiritual support, and they prayed to God always with their family and in religious institutions to increase their social activities and their social contributions. The following quotes reveal such type of coping mechanisms:

> *“I used holy water in a religious place. It has some changes for the time being. Since I developed the disease, I have used holy water at Sodo Town [Kokate] Church (Gedam) and other churches. I believe holy water helps and heals me, but it requires strong belief and courage.”* (IDI, 8)

> *“My prayers with my family every night helped me in my life. The belief that there is a God gives me hope and encouragement. God has created man to live and die, but with a purpose. So, I believe he has a purpose for me and a reason for me to have this condition. So, I believe that he has helped me accept my condition and shape my beliefs about death. Reading the word of God is so helpful, and it provides a sense of hope in my life.”* (IDI, 5)

#### Modern medicine and maintaining hygiene as a coping mechanism

According to participant’s responses, seeking medical treatment from a health facility and maintaining hygiene help participants cope with physical health challenges, social, and psychological health challenges. Participants also described receiving medical support and information as helpful for managing their stress related to psychosocial problems they encounter in their day-to-day activities. Participants discussed that their physical health problem were relieved when they kept their hygiene clean and washed their feet with hot water and soap. It is also critical to follow the advice of health professionals to manage this tedious illness. This notion is well described as follows;

> *“I have been following the health professionals’ guide, and now, praise God, I am healthy and my feet are improved. When I keep my feet clean and wash my feet with hot water, the pain and swelling get relieved. After receiving treatment, other people suffering from this disease were surprised to learn about my condition. Now I teach other Podoconiosis patients to seek care from Mossy Foot International.”* (IDI, 7)

#### Social support

Participants reported that they sought social support from their family and community, which is both practical and emotional. Some of the participants reported support from society and family members such as parents, peers, neighbors, and health professionals. This strategy involved getting help with physical, social, and psychological problems from parents and maintaining good relationships with close friends to ensure support. Support from family members and society is an important and healthy strategy required to mitigate their hurtful experiences related to their illness. This result is well explained as follows;

> *“My family and I get along well socially, and I’m delighted for them. When I was incapacitated, my wife did her best to take care of me by preparing traditional medicines and home remedies. The majority of my neighbors are friendly to me; they don’t tease me but instead help me. I am an active member of neighborhood organizations, which benefits our relationship. I don’t recall any negative attitudes toward my illness from either Edir or Equb community organizations. My community organizations make me happy.”* (IDI, 13)

#### Wearing thick clothes and shoes as a coping strategy

Study participants hide the swelling of their lower leg from community members by wearing thick clothes and shoes. Participants discussed that they hide swelling of their lower leg during social activities like weeding and funerals. Participants reported that they try to conceal themselves so that others do not recognize them and no stigma and discrimination will occur to them. To cope with stigma and discrimination participants wear thick clothes and shoes. This notion is well explained as follows;

> *“To cope with this disease, I used to wear wide and very long clothes when I wanted to go out. So, people can’t identify me easily. Even when I went to the clinic, if people asked me where I was going, I told them I was going to ask the patient over here. I wear shoes, too.”* (IDI, 7)

#### Traditional medicines and plants as coping mechanisms

Traditional medicines and plants have been used as coping mechanisms for Podoconiosis. These practices are deeply rooted in cultural and historical contexts, with indigenous communities often relying on traditional healing methods to address physical, mental, and emotional well-being. The use of traditional medicines and plants as coping mechanisms is based on the belief that nature provides remedies for a wide range of ailments, and these practices are often intertwined with spiritual and holistic approaches to health. Study participants reported traditional medicine/herbal treatment and plants as treatment options to cope up from physical health problems.

A participant explained as follows;

> *“I used a variety of herbal remedies and plants to treat myself. I had treatment using a variety of leaves, including coffee tree leaves, which are commonly used to make coffee in the area. The leaves of coffee rubbed on the feet and the water in the coffee leaves dropped on the feet. There are folk healers who are familiar with the leaves.”* (IDI, 14)

## Discussion

The study found that participants predominantly perceived witchcraft or sorcery as the main cause of Podoconiosis, contrasting with findings from Rwanda, where soil contact was commonly cited, likely due to socio-demographic differences and the involvement of health professionals in the prior study (29). Similarly, research from Northern Ethiopia reported misconceptions such as Podoconiosis being caused by standing up too fast, hot soil burning the skin, blood contact, evil spirits, or divine curses (30). However scientific findings revealed that Podoconiosis is caused by longstanding exposure to red clay soil of volcanic origin (2). This result indicates that most participants failed to notice the cause of Podoconiosis. This may cause a delay in seeking health care, thus aggravating the condition and reducing the chance of recovery and making patients bedridden for a long period of time. This might be due to participant’s low literacy level, and or communities’ negative perception towards to Podoconiosis which might have an influence on the understanding of Podoconiosis and its treatment options.

The current study also showed that the cause of Podoconiosis is related with hereditary/familial. The participants rarely mentioned heredity as the cause of Podoconiosis. This finding is consistent with a study conducted in Ethiopia where, people believe that anyone who has the disease in the bloodline of the family can pass it on to the children born to that person (31).

In the present study, some participants reported that barefoot exposure to poison and worms was a perceived cause of Podoconiosis. Though the term soil or dust was rarely mentioned, participants referred it to poison and other harmful things in the soil. This study is corroborated by a previous study conducted in Rwanda and Southern Ethiopia. (29,32).

According to the current study Podoconiosis patients had problems of interacting with community members. This resulted in patient’s isolation and withdrawal from society, avoiding social gatherings activities and had problems of interaction even within their own families. This finding is in line with study conducted in Southern Ethiopia and Northern Ethiopia where the study reported that participants had experienced one or more forms of social stigmatization at school, church, or in the market place including school dropout, forced exclusion, not buying products from them, shunning, pointing at them, nose pinching and insulting (33,34). This result indicates that there is maltreatment and negative attitude/perception in the community toward people living with Podoconiosis.

According to the responses the participants experienced psychological health problems due to Podoconiosis. Most of the study participants reported experiencing occasional feelings of sadness or depression. This result is also reported similarly in other studies conducted in Ethiopia, Cameroon and Rwanda. (16,35,36). This finding indicates that Podoconiosis often causes swelling, pain, itching, and discomfort in the lower limbs. These physical symptoms can be chronic and severe, leading to decreased mobility and a decreased quality of life. Constant pain and discomfort can contribute to feelings of depression and sadness. Individuals with visible symptoms of Podoconiosis may face social stigma and discrimination. This can result in social isolation, which can be emotionally distressing and lead to feelings of loneliness and depression. This calls a need to provide physical therapy and psychosocial support to people living with Podoconiosis.

The majority of participants in this study experienced feelings of hopelessness, sadness, shame and inferiority associated with disfiguring look of their feet and bad odor which were aggravated during acute attacks. A similar study in Ethiopia reported that people living with lymphedema (Podoconiosis) had experienced anger, sadness, and shame associated with a lack of support from their relatives, family members, and community members and some of the Podoconiosis patients reported experiencing suicide ideation/thoughts, feeling helpless, and feeling abandoned (36,37). This finding indicated that people living with Podoconiosis need special care and support from family and community members.

The present study also found that almost all participants reported financial problems and loss of livelihood due to the illness. The majority of participants suffering from Podoconiosis said they were not able to work as before because of their condition. This finding is consistent with those of other studies conducted in Ethiopia (37–39). This finding highlights the significant economic impact of illness on individuals and their families. This can lead to increased financial stress, which may further exacerbate the health condition of the individual. The findings suggest the need for increased social support systems to help individuals cope with the financial consequences of illness. This could involve government-funded programs, non-profit organizations, or even private initiatives aimed at providing financial assistance to people living with Podoconiosis.

The current study revealed that the majority of the study participants used home remedies as well as traditional medicine for treatment. Participants believed that the illness had no specific treatment and they preferred home remedies and traditional medicine. This result is not similar with previous studies where participants used an unidentified herb named “Shingung” in Amharic. Some herbs were unavailable during the dry season, and participants reported concerns about addiction and withdrawal symptoms, especially with Shingung (40). This difference might be due to socio-cultural variation among participants. This finding indicates that local illness treatment makes the people living with Podoconiosis perceive that illnesses have no medical treatment from health facilities, and rely on the traditional medicines rather than seeking care from health facilities; leading to the development of misconception about treatment options.

According to current the study, unavailability of the services in nearby health centers is the most common barrier to seeking care which was predominantly mentioned by participants. Participants reported that they went to their nearby health center to seek care but the unavailability of services in the center prevented participants from accessing services. This result is consistent with a study conducted in Northern Ethiopia, where a lack of adequate healthcare provision and unavailability of treatment in health centers and health facilities were reported as the barriers to seeking care (41). This finding indicates that Governmental and Non-governmental organizations have paid little attention to Podoconiosis. Due to the unavailability of services in the district, patients may experience delays in receiving necessary treatments, tests, or medications. This can lead to worsening health conditions, increased pain and discomfort, and in severe cases, even life-threatening situations. The inability to access essential services can also result in increased stress and anxiety for patients and their families.

The present study showed that Podoconiosis patients seek social support from their family and community which are both practical and emotional. Some of the participants reported support from society and family members such as parents, peers, neighbors and health professionals. This result is not similar to study conducted in Ethiopia where Some unmarried Podoconiosis patients participated in the study, and said they would avoid marrying unaffected people (18).

### Strengths and Limitations of the study

This study’s strengths lie in its home-to-home visits, which provided insights into the real-life experiences of individuals living with Podoconiosis, including both those who had and had not attended health facilities. The research employed an open, emerging approach, with data collection, analysis, and write-up guided by ideas derived directly from the data rather than preconceived notions. Additionally, face-to-face interactions allowed for capturing important non-verbal cues. However, the study faced limitations such as potential deviations in meaning during translation of data into English, limited related literature for in-depth discussion, and possible influence of social desirability bias on participants’ responses.

### Implications of the result

The study revealed widespread misconceptions among the community regarding the causes of Podoconiosis, leading to stigma and mistreatment of affected patients. Podoconiosis patients already endure significant physical, social, and psychological challenges, which are exacerbated by this discrimination, highlighting that the disease is both neglected and affects marginalized populations. The findings emphasize the need for health care providers, managers, and stakeholders in Southern Ethiopia to implement health education and behavior change strategies to raise awareness and reduce stigma. The lack of accessible health services has contributed to delays in seeking care, indicating limited attention from both governmental and non-governmental organizations. Strengthening local health infrastructure is crucial to address these gaps. Furthermore, the study provides baseline data for future research, recommending quantitative surveys and engagement with health workers and policymakers to better understand and address Podoconiosis in Ethiopia.

## Conclusion

The study participants believed that Podoconiosis is caused by witchcraft, heredity, walking barefoot, or curses from God or evil, which negatively affects patients’ health-seeking behaviors for prevention and treatment. Podoconiosis leads to severe physical, economic, psychological, and social consequences, including social isolation, frequent divorce, and lack of support from family and community. As a result, patients often experience hopelessness and suicidal thoughts. Providing love, care, and social support can significantly improve the quality of life of those affected.

## Data Availability

All data produced in the present study are available upon reasonable request to the authors

https://www.NTDs.com

## Ethics approval and consent to participate

Ethical approval was obtained from Jimma University Research Ethical Review Board, Ethiopia. Permission paper was obtained from the Wolaita zone Health department and Sodo Zuriya health office. The right of research participants was maintained by ensuring non-maleficence and underscoring the benefits of the study. Study participants were informed adequately about the purpose of the study, voluntary participation, and the right to participate or withdraw at any time. To ensure their privacy and autonomy, code was given to participants and informed as the study uses this code in place of their names in connection to the study findings or their answers on discussions or interviews. Time was given to them to reflect and provide a detailed explanation of the issue. Individual-based informed consent was obtained and a separate consent was also obtained for audio-recording and the consent taken was also included.

## Acknowledgements

We would like to acknowledge Jimma University, Faculty of Public health for assistance of undertaking this research. We would also like to express our gratitude to study participants, Wolaita Zone health department health care workers.

## Authors’ contributions

MM: Conceptualization, Data curation, Formal analysis, Investigation, Methodology, Project administration, Resources, Software, Validation, Visualization, Writing – original draft, Writing – review & editing. ET: Investigation, Software, Validation, Visualization, Writing – review & editing. FN: Supervision, Validation, Writing – review & editing. YK: Supervision, Validation, Writing – review & editing.

## Funding

No funding was obtained for this study.

## Availability of data and materials

Data can be available from the corresponding author upon reasonable request.

## Competing interests

The authors declare there is no competing interest among the authors.

## Notes

### Competing Interest Statement

The authors have declared no competing interest.

### Funding Statement

This study did not receive any funding

### Author Declarations

Ethical approval was obtained from Jimma University Research Ethical Review Board, Ethiopia.

